# Scalable RT-LAMP-based SARS-CoV-2 testing for infection surveillance with applications in pandemic control

**DOI:** 10.1101/2022.06.27.22276704

**Authors:** Dan Lou, Matthias Meurer, Svetlana Ovchinnikova, Robin Burk, Anna Denzler, Konrad Herbst, Ioannis A. Papaioannou, Yuanqiang Duan, Max L. Jacobs, Victoria Witte, Daniel Ürge, Daniel Kirrmaier, Michelle Krogemann, Krisztina Gubicza, Kathleen Boerner, Christian Bundschuh, Niklas M. Weidner, Uta Merle, Britta Knorr, Andreas Welker, Claudia M. Denkinger, Paul Schnitzler, Hans-Georg Kräusslich, Viet Loan Dao Thi, Andreas Deckert, Simon Anders, Michael Knop

## Abstract

Throughout the current SARS-CoV-2 pandemic, limited diagnostic testing capacity prevented sentinel testing of the population, demonstrating the need for novel testing strategies and infrastructures. Here, we describe the set-up of an alternative testing platform, which allows scalable surveillance testing as an acute pandemic response tool and for pandemic preparedness purposes, exemplified by SARS-CoV-2 diagnostics in an academic environment. The testing strategy involves self-sampling based on gargling saline, pseudonymized sample handling, automated 96-well plate-based RNA extraction, and viral RNA detection using a semi-quantitative multiplexed colorimetric reverse transcription loop-mediated isothermal amplification (RT-LAMP) assay with an analytical sensitivity comparable to RT-quantitative polymerase chain reaction (RT-qPCR). We provide standard operating procedures and an integrated software solution for all workflows, including sample logistics, LAMP assay analysis by colorimetry or by sequencing (LAMP-seq), and communication of results to participants and the health authorities. Using large sample sets including longitudinal sample series we evaluated factors affecting the viral load and the stability of gargling samples as well as the diagnostic sensitivity of the RT-LAMP assay. We performed >35,000 tests during the pandemic, with an average turnover time of fewer than 6 hours from sample arrival at the test station to result announcement. Altogether, our work provides a blueprint for fast, sensitive, scalable, cost- and labor-efficient RT-LAMP diagnostics. As RT-LAMP-based testing requires advanced, but non-specialized laboratory equipment, it is independent of potentially limiting clinical diagnostics supply chains.

**One-sentence summary:** A blueprint for scalable RT-LAMP test capacity for the sensitive detection of viral genomes demonstrated by SARS-CoV-2 surveillance testing.

## INTRODUCTION

In a global pandemic such as the current with the severe acute respiratory syndrome coronavirus 2 (SARS-CoV-2), the development of rapid and sensitive diagnostic techniques for the identification of infected individuals is of highest priority to track infection chains and support pathogen containment. This applies especially to infectious diseases with ambiguous symptoms and an asymptomatic infectious period, such as the coronavirus disease 2019 (COVID-19) (1–6) or the acquired immunodeficiency syndrome (AIDS) (7). To evaluate and respond to a potential pathogen threat, nucleic acid-based diagnostic methods are ideal since the required reagents can be rapidly created and validated, provided that genomic sequence information of the pathogen is available. In the SARS-CoV-2 pandemic, the early availability of the genome (8) allowed the development of reliable reverse transcription-quantitative polymerase chain reaction (RT-qPCR) primer sets in Germany (9) and many other countries before the virus started to spread. Besides the need for specific probes, RT-qPCR-based testing capacity is also dependent on preexisting diagnostics infrastructure including highly specialized equipment for sample collection, processing, and analysis. Throughout all waves of the pandemic, the diagnostic demand exceeded the production capacities of the device manufacturers and led to a global shortage of RT-qPCR equipment severely limiting testing capacity. Insufficient testing capacity is considered a tipping point between controlled and uncontrolled viral spread (10). In Germany, the initial RT-qPCR testing capacity increased within weeks to reach a capacity of more than 800,000 tests per week in mid-April 2020 - two weeks after the peak of the first wave (11,12). This was by far not enough to satisfy the need, as indicated by the backlog of samples for testing during different phases of the pandemic (11,12) or the fraction of positively tested samples, which is considered another indicator for testing capacity. Inadequate testing means that many infected individuals go undetected (13). This demonstrates that the buildup of RT-qPCR testing capacity during the entire SARS-CoV-2 pandemic was never sufficient for applications other than passive surveillance (based on symptoms) and contact tracing (by health authorities). Starting in the third quarter of 2020, rapid SARS-CoV-2 antigen tests were made available aiding active viral containment. As the development of these tests is laborious and time-consuming, they are not suitable as an early response tool. Additionally, considering their limited accuracy and sensitivity (14,15), antigen tests are inferior compared to nucleic acid amplification-based pathogen detection techniques. A high false-positive rate for some of the many different brands (16) (https://ec.europa.eu/health/system/files/2022-05/covid-19_rat_common-list_en.pdf) requires additional nucleic acid testing to obtain a valid diagnostic result, making them a (lucrative) addition - rather than a competitor - in the overall test offering.

The limitation in RT-qPCR test capacity prompted the molecular biology research community to seek independent and affordable alternatives for sensitive pathogen detection. Very early in the pandemic loop-mediated isothermal amplification (LAMP) (17,18) coupled with a reverse transcriptase (RT-LAMP) was developed and validated as an alternative viral genome detection method (19*,20) (*first time posted on MedRxiv on February 29, 2020). RT-LAMP is a particularly attractive substitute for RT-qPCR because it does not depend on the availability of specialized equipment such as thermocyclers. The reagents are easily accessible or can be largely self-produced. In addition, the RT-LAMP patents and the patents for the enzymes needed for the RT-LAMP reaction have expired (21). Conventional nucleic acid amplification methods for viral detection include a fluorescence readout. In RT-LAMP, this can be replaced by using fluorescence dyes to detect the produced DNA, or dyes to detect a chemical change associated with a successful amplification, e.g. a pH drop (22) or the formation of a magnesium pyrophosphate precipitate (23). In addition, since fluorescence and color readout can constitute bottlenecks, we and others developed DNA barcoding strategies to permit pooled analysis of many samples using next-generation sequencing (termed LAMP-sequencing) (24–26), providing the possibility of a massively scaled up sample throughput.

We finished validation of the method within a short period (24–27) and started using the colorimetric RT-LAMP as an alternative to commercial RT-qPCR testing capacity for sentinel studies and surveillance testing of larger groups of people. Using this new testing infrastructure, we avoided putting an additional burden on the locally available diagnostic capacity or affecting the supply chains of RT-qPCR diagnostics. This was important to justify sentinel testing in a situation where test capacity available for contact tracing by health authorities was strongly limited. In this publication, we describe the implementation of a high-throughput RT-LAMP-based test station including all steps from sampling to result announcement. We describe procedures for self-sampling using an optimized gargling protocol that integrates into a simple workflow for robotic processing of the samples and improved the RT-LAMP assay for enhanced robustness resulting in sensitivity and specificity values comparable to RT-qPCR. Together with detailed standard operating procedures and open-source software solutions for sample registration and tracking as well as RT-LAMP time course measurements, we report a high-capacity test station that is based on research-grade equipment largely independent of supply chains for clinical diagnostics. We demonstrated the application of the test platform in the context of the current pandemic by testing more than 35,000 samples for a SARS-CoV-2 infection over nine months.

## RESULTS

### Gargle sampling procedure and sample stability

Conventional nucleic acid testing for SARS-CoV-2 diagnosis is based on pharyngeal swabs (naso-/oropharyngeal (NP/ OP) swabs). This limits the scalability of testing of large populations due to a major bottleneck constituted by the requirement of trained personnel for swabbing. Saliva and gargle samples, as alternatives to NP/ OP swab specimens, can be easily self-collected and provide equivalent sensitivity for SARS-CoV-2 diagnostics (28–30). To account for simple sample processing in the test station, we developed a specialized gargling kit as well as a 60 s protocol for self-collection of a combined fraction of gargling liquid, saliva, expectorated cough, and nasal mucus tailored to the kit. The kit contains a small bottle with 5 ml of gargling liquid (0.9% saline) and a straw that allows, after gargling, easy return of the liquid from the mouth into the plastic bottle. The pointy tip of the bottle is used to dispense ∼1 ml of the gargling fluid into a barcoded Micronic tube (Micronic, Lelysta, Netherlands), which is returned to the test station for analysis (Figure 1A). The Micronic tube can be processed directly by standard liquid handling platforms, thus avoiding the need for manual pipetting steps in the laboratory and thereby decreasing human error as well as infection and contamination risks.

**Fig. 1:**
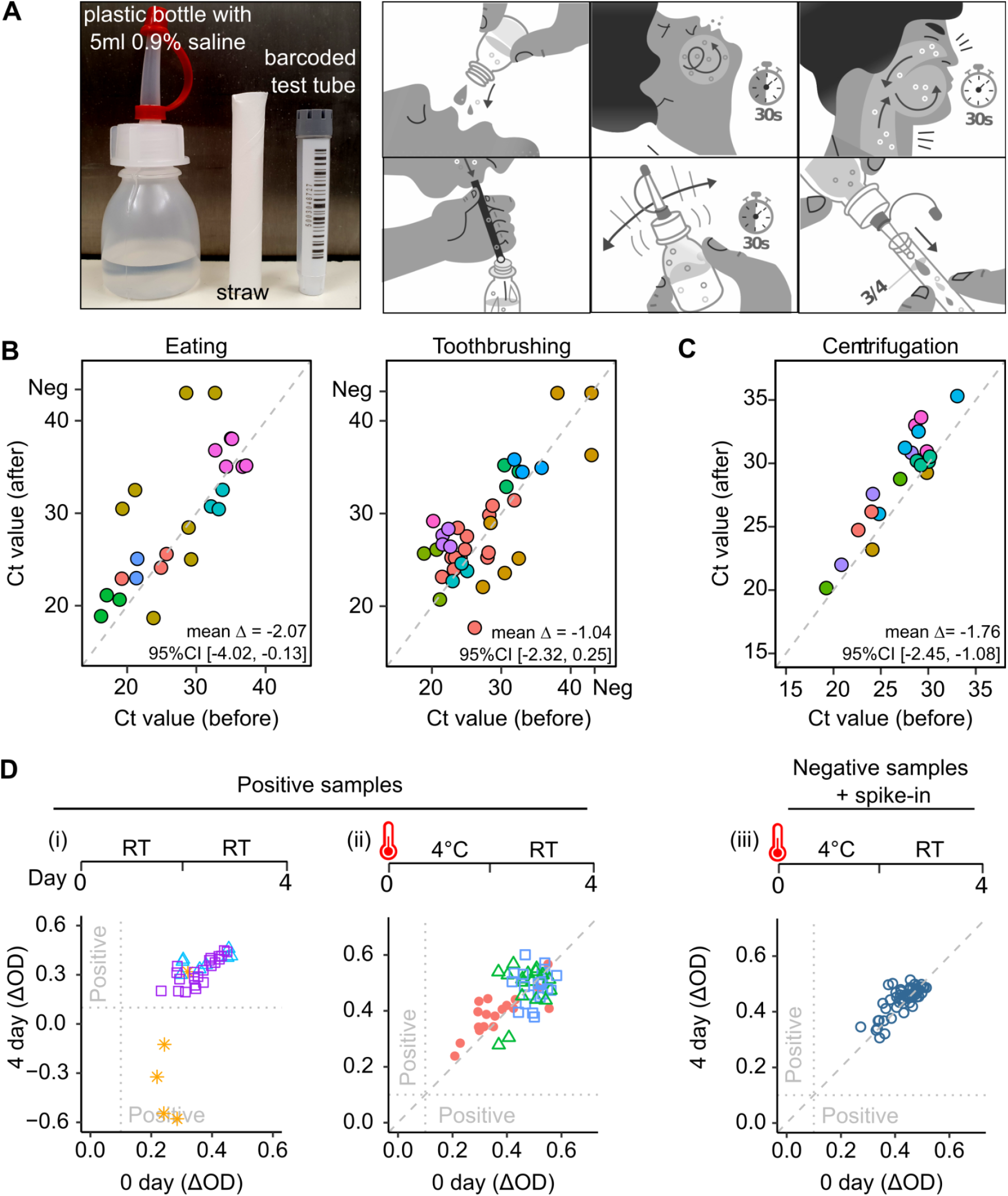
Development of a gargling self-sampling procedure for SARS-CoV-2 detection by RT-LAMP. (**A**) Test kits for self-sampling contain a plastic bottle (food grade) with 5 ml sterile 0.9 % saline for gargling, a straw (food grade), and a barcoded test tube. To collect the sample, participants gargle with the saline for 30 s and may also collect expectorated cough and nasal mucus for additional 30 s. Subsequently, the sample is transferred into the plastic bottle using the straw. The sample is homogenized by shaking for 30 s. After cutting off the tip of the bottle, the sample is transferred into the barcoded test sample tube, which is closed tightly and sent for analysis by RT-LAMP. (**B**) Longitudinal samples collected before and after specific behaviors of test subjects (eating, toothbrushing) were analyzed by RT-qPCR. (**C**) Each longitudinal sample was divided into two aliquots. One aliquot was centrifuged at 3500 g for 1 min and the supernatant was taken for RT-qPCR analysis. The other aliquot was mixed well by vortex and analyzed directly by RT-qPCR. (**D**) Longitudinal sample series from different patients were analyzed by RT-LAMP. Colors and shapes indicate individual patients. (i) Each sample was divided into two aliquots. One aliquot was immediately frozen at -20 °C while the other was kept at room temperature (RT) for four days before being analyzed by RT-LAMP. (ii) Frozen aliquots of longitudinal sample series were thawed and heat-inactivated (>85 °C for 5 min). The aliquots were split and one half was frozen again while the other half was kept for two days at 4 °C and two days at room temperature prior to RT-LAMP analysis. (iii) Similar to (ii), but using negative samples and spike-in of a highly positive sample. (**B-D**) Colors/ symbols indicate samples from individual patients. Several samples per patient were used for each comparison.

To test the sampling strategy, several voluntary test subjects with an acute SARS-CoV-2 infection were recruited. Multiple longitudinal samples were collected during their infections (to up to 15 days after diagnosis, Supplementary Figure S1), and the exact time points of sample collection and factors potentially affecting viral loads in the samples were recorded. This resulted in longitudinal sample series of at minimum 15 up to 143 samples per person covering in total a cycle threshold (Ct) ranging from ∼Ct15 to >Ct35 (Supplementary Figure S1). We compared Ct values before and after specific behavior of the test subjects or sample handling to judge the impact on SARS-CoV-2 diagnosis (Figure 1B-D). This revealed an increase of the Ct values by approximately two cycles for food intake (averaged Δ(Ct[after]-Ct[before]) = -2.07; 95% confidence interval (CI) [-4.02, -0.13]; *p*-value = 0.04). For tooth brushing (sample taken earliest 10 min after teeth brushing) this effect was less prominent (Δ = -1.04; 95% CI [-2.32, 0.25]; *p*-value = 0.11), but also tended to reduce the viral load in the sample. Based on these observations we recommended collecting the gargle sample in the morning before breakfast or at least 60 min after the last food uptake. Furthermore, we observed that centrifugation of the sample (3500g for 1 min) increased the Ct values on average by almost two (Δ = -1.76; 95% CI [-2.45, -1.08]; *p*-value = 3*10^-5), indicating that a large amount of the virus is present in larger particles or infected cells that can be easily sedimented (Figure 1C). To evaluate sample stability and viral genome detection under different conditions, some samples were divided into two fractions. One fraction was immediately frozen at -20 °C, while the other tube was kept at room temperature (RT) for four days before it was frozen. Subsequently, RT-LAMP analysis was performed side-by-side for these samples (Figure 1D (i)). For the majority of samples, the RT-LAMP tested positive for both fractions. For one individual, we observed four samples with a low viral load (Ct>30) tested negative in the LAMP assay after four days of storage at RT. For the sake of safety and convenience, a heat inactivation step (5 min at 85-90 °C) was included in the sample processing procedure (31). We observed that the heat inactivation step and subsequent storage of the samples at 4°C (for later validation of positive RT-LAMP results) did not negatively influence detectable viral load or sample stability (Figure 1D (ii; iii)). Together these experiments suggested a convenient and robust sample collection and handling protocol for SARS-CoV-2 diagnostics.

### Implementation of high throughput RT-LAMP diagnostics based on a colorimetric readout

To accommodate high numbers of samples, we implemented and validated an automated RNA extraction with a laboratory liquid handling device (Hamilton NGS STAR), which resulted in a time saving of ∼55 min (40%) for one sample plate at similar quality (Supplementary Figure S2, SOP1). In addition, we implemented a high throughput RT-LAMP assay in a multiwell format (Figure 2, SOP2). The Micronic tubes containing the gargle samples were compatible with 96-position racks, which could directly be loaded onto the Hamilton platform. Per 96-position rack, 92 Micronic tubes with test samples and four controls were included (Figure 2A). Three positive controls with known concentrations (e.g. Ct=29, 32, 36) were used to evaluate the performance of RNA isolation and RT-LAMP assay. In addition, a negative control was incorporated.

**Fig. 2:**
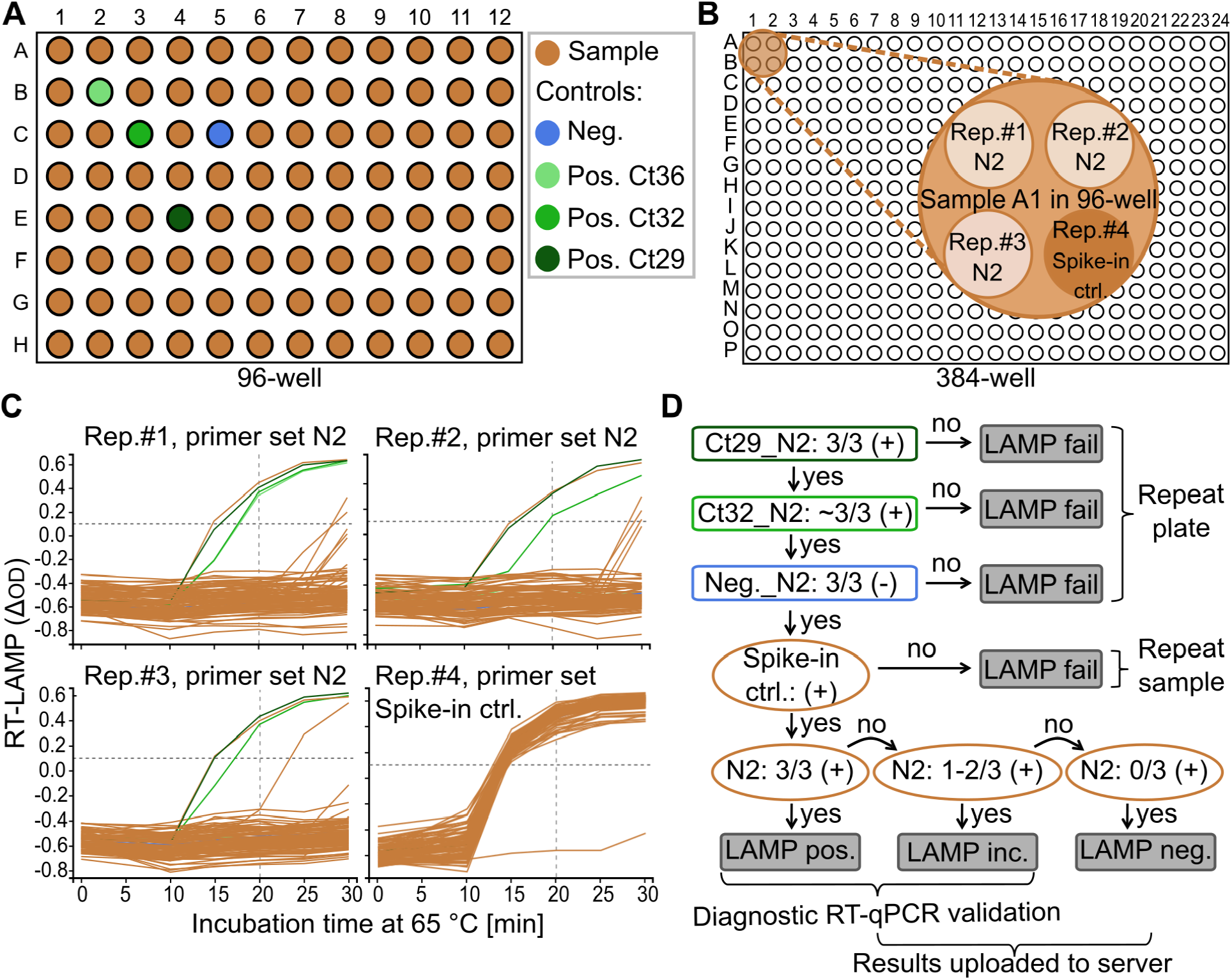
Optimized RT-LAMP format and analysis of data. (**A**) The layout of a 96-well plate with 92 samples and four controls. Positions of the controls were assigned at random. (**B**) For RT-LAMP analysis, four times 5 μl of the RNA were dispensed into a 384-well plate and the RT-LAMP assay was conducted using either three replicas of the N2 primer (Rep. #1-3, N2) or one replica of a positive spike-in control primer (Rep. #4, spike-in ctrl.) as indicated. (**C**) Time-course analysis of the color change (ΔOD) of the LAMP reaction. Controls as indicated in (A). (**D**) Annotation of the decision matrix. (+) indicates ΔOD ≧0.1 at 20 min; (-) indicates ΔOD < 0.1 at 20 min. LAMP positive (3/3 positive replicates) and LAMP weakly positive (1/3 or 2/3 positive replicates) results were validated by RT-qPCR.

The sensitivity of detection of an RNA template in an RT-LAMP reaction is dependent on many factors including the used primer set or combinations of primer sets (19,32,33). Moreover, in some samples, in particular samples with low concentrations of isolated nucleic acids, the initiation of the RT-LAMP reaction can lead to spurious amplification impairing RT-LAMP specificity (24,34). We used well-established primer sets targeting the SARS-CoV-2 N gene (N2) (32) alone or in combination with a primer set targeting the SARS-CoV-2 Orf1b gene (N2/ Orf1b-2) (33). In addition, we converted our previous 96-well-based RT-LAMP assay format (24) into a 384-well assay format, containing three replicates per sample to detect SARS-CoV-2. A fourth reaction included a primer set for the detection of an internal control RNA spiked into the lysis buffer used during the RNA extraction (Figure 2B). Integration of an internal control RNA (i.e. from Zika virus) in amounts close to the detection limit using the corresponding RT-LAMP primer set (Zika2-5 from (35)) provides a measure to assess the integrity and sensitivity of the whole RNA isolation and detection procedure. To quantify the RT-LAMP reaction, the colorimetric changes (from red to yellow) were measured every 5 min using a microplate reader, as described before (24). To monitor RT-LAMP amplification of target sequences, the differences in absorbance (ΔOD) of each well (replicate) at 434 nm and 560 nm were plotted against the incubation time at 65°C (Figure 2C). For automated annotation of RT-LAMP results, we established exact criteria to decide on the diagnostic status for each sample, as outlined in the decision tree for annotation of the results in Figure 2D. This was implemented in an open-source tool termed “LAMP plate viewer” (https://github.com/anders-biostat/lamp_plate_analysis) that inputs files from the plate reader as well as the documented positions of the Micronic sample tubes and associated barcodes. The test station personnel evaluated the RT-LAMP plates based on the decision tree and assigned a status for each sample assisted by the automated pre-classification of the “LAMP plate viewer tool”. After verification of the assignments by the test station personnel, the diagnostic results were directly uploaded to a database, where test participants could retrieve their individual results via a webpage. Taken together, we implemented a convenient high throughput sample analysis pipeline allowing quick result visualization, automated pre-classification of the results, and manual quality control for reliable SARS-CoV-2 diagnostics.

### Diagnostic sensitivity of the colorimetric RT-LAMP testing platform

To validate the performance of the colorimetric RT-LAMP assay, we compared the results to the results obtained using RT-qPCR for a total of 819 gargle samples obtained from both longitudinal sample series (347 samples distributed on 8 plates) and regular testing (472 samples distributed on 7 plates) on different days. After RNA extraction, both assays were performed within 24 to 48 hours (RNA samples were stored at -20°C in between). For the RT-LAMP assay, 5 μl of RNA was used for each replicate per sample, whereas for the RT-qPCR 10 μl was used for a single reaction per sample. For RT-LAMP, either the primer set N2 or a mixture of the N2/ORF1b-2 primer sets were used, while the RT-qPCR assay used primers for the E gene (TIB MolBiol LightMix Modular Sarbecovirus SARS-CoV-2, Cat.-no. 50-0776-96) that bind to regions that do not overlap with the regions amplified by RT-LAMP. For the RT-LAMP assay, samples with at least one replicate exhibiting a ΔOD>0.1 at 20 min after the start of the incubation at 65 °C were considered positive. For the RT-qPCR, we strictly followed the instructions of the manufacturer, including color compensation and the acquisition of melting curves to decide for ambiguous RT-qPCR curves whether a sample was positive or negative (see Materials and Methods). For visualization, the mean ΔOD value at 20 min after the start of the incubation at 65 °C of the positive replicates in the RT-LAMP assay was plotted against the corresponding Ct value in the RT-qPCR assay (Figure 3A). In total, five samples that were tested positive by RT-LAMP were negative by RT-qPCR. Since all these samples were derived from longitudinal sample series from different diagnosed COVID-19 patients, we consider these as false-negative RT-qPCR samples. We also observed four samples that were RT-LAMP negative but RT-qPCR positive samples; including one sample from the longitudinal sample series and three from regular testing, all with CT>35. We consider these samples as false-negative RT-LAMP samples (Figure 3A). Taken together, in our implementation of automated SARS-CoV-2 testing all samples with a diagnostic Ct value of Ct34 and larger contained at least one replicate that scored positive in the RT-LAMP assay (285 of 285), whereas 83.3% of the samples with Ct values between 34 and 36 were tested positive with at least one replicate in the RT-LAMP assay (20 of 24). The overall specificity of the RT-LAMP assay with this set of samples was 99.0% (95% confidence interval: 97.6% - 99.6%) (Figure e 3B, Table S1).

**Fig. 3:**
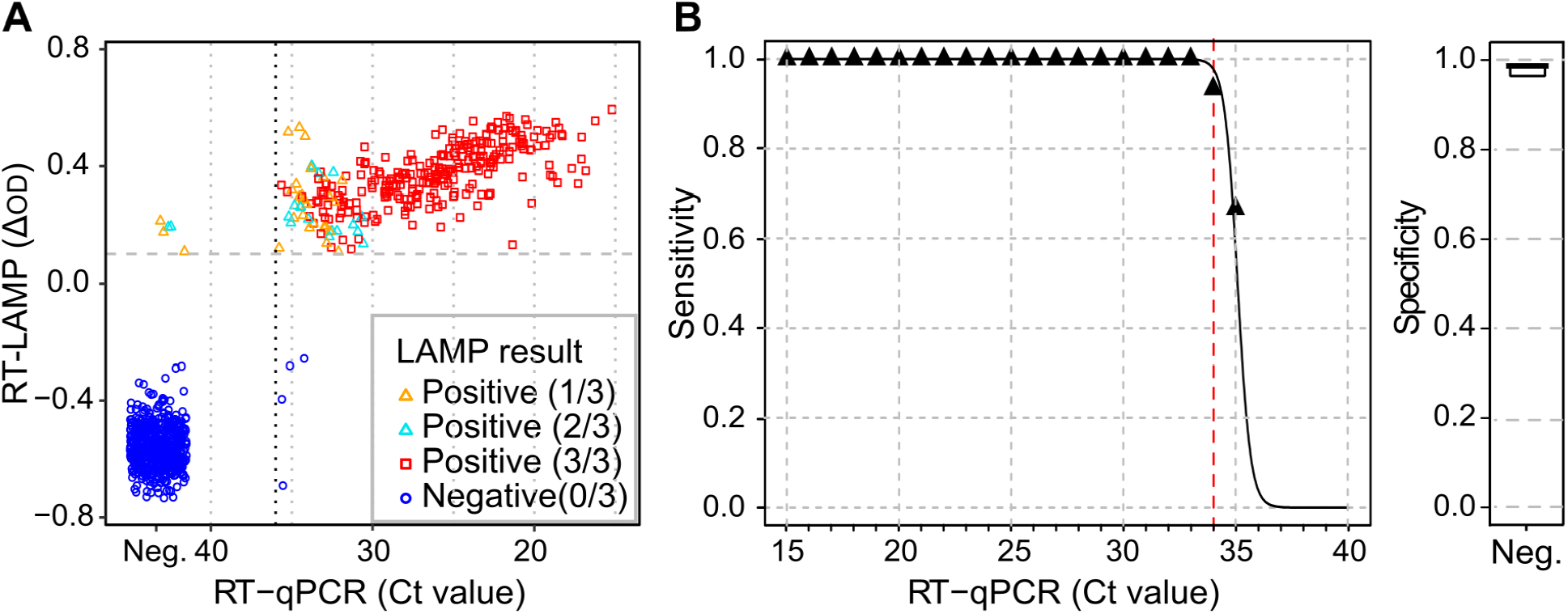
Specificity and sensitivity of the optimized RT-LAMP assay. (**A**) Scatter plot showing a comparison of RT-qPCR (Ct values) and RT-LAMP results (ΔOD values at 20 min after the start of the 65 °C incubation) of RNA samples. Color and symbol codes indicate the number of positive replicates for the RT-LAMP analysis. (**B**) Sensitivity (left) and specificity (right) of the RT-LAMP assay (derived from data in (A) and Table S1).

### Data handling

We developed an open-source software solution to handle sample logistics, laboratory information flow, communication of results, and further administrative tasks. A database kept track of all samples and received test results directly from the “LAMP plate viewer” tool. Participants could query their test results by entering an access code that was provided on a paper slip in the self-sampling test kit. The staff of the testing station could retrieve detailed information on the history of each sample (Supplementary Figure S4), as well as perform database queries for statistics and, if needed, invoicing. The software is written in Python, using the Django framework, and available for download from https://github.com/anders-biostat/covid-test-web-site. Care has been taken to allow for easy customization and integration. In fact, when another institution decided to copy our testing set-up, their IT staff succeeded in setting up and customizing our software using only the published documentation, i.e., without the need to consult with us - thus demonstrating that the platform is mature, versatile, well documented and easily deployable.

The European Union has a high level of employee protection and data privacy is considered extremely important. This is governed by the General Data Protection Regulation in the European Union and the European Economic Area. Therefore, special care was taken to safeguard data privacy: All samples were identified only by barcode and access code, i.e., fully anonymized. However, as German law requires diagnostic laboratories to notify health authorities of positive results of tests on certain pathogens, including SARS-CoV-2, the participants’ identities had to be queried before revealing any test results. To obtain optimal data security, the server encrypted these data immediately upon receipt using the RSA public-key cryptography scheme, and the private key needed to decrypt the data was given to the local health authorities only, making it impossible for anybody else to access any personal data. Only in the case of a positive test result, the personal data of the corresponding test participant was decrypted by local health authorities to initiate appropriate measures such as contact tracing. This data encryption strategy increased data security and made it impossible to link test results to the personal information of the test participants for people who possibly gained accidentally or with malicious intent access to the database.

Having the diagnostic assay as well as the IT infrastructure at hand, we were able to offer SARS-CoV-2 surveillance testing in the academic environment of the University of Heidelberg to support local viral containment. To be able to face the challenge of sample logistics in campus-wide testing, we developed a system that can be easily scaled up to collect sample tubes from large groups of individuals (Figure 4A): The system included a central distribution point for test kits with daily opening hours to handout test kits in defined quantities to representatives of institutes or workgroups on the campus, which handed over the test kits to individual test participants. Participants performed the gargle self-sampling procedure at home and registered their samples on the test station server with a 12-digit access code associated with the individual barcode on the Micronic tube. Permanently installed sample collection boxes were set up at central locations where test participants could drop off their samples. In addition, small, mobile collection boxes were distributed on request allowing individual institutes, work groups, or organizers of university events to collect samples in a decentralized manner and return them to the test station. Workflows set up in the test station permitted the analysis of returned samples in batches of up to 8×92 samples within 5-6 hours until the results were published on the test station server (Figure 4B). Participants retrieved their results via the individual access codes. In case of positive RT-LAMP results, health authorities were automatically informed and deciphered the corresponding contact information using the private key (Figure 4A).

**Fig. 4:**
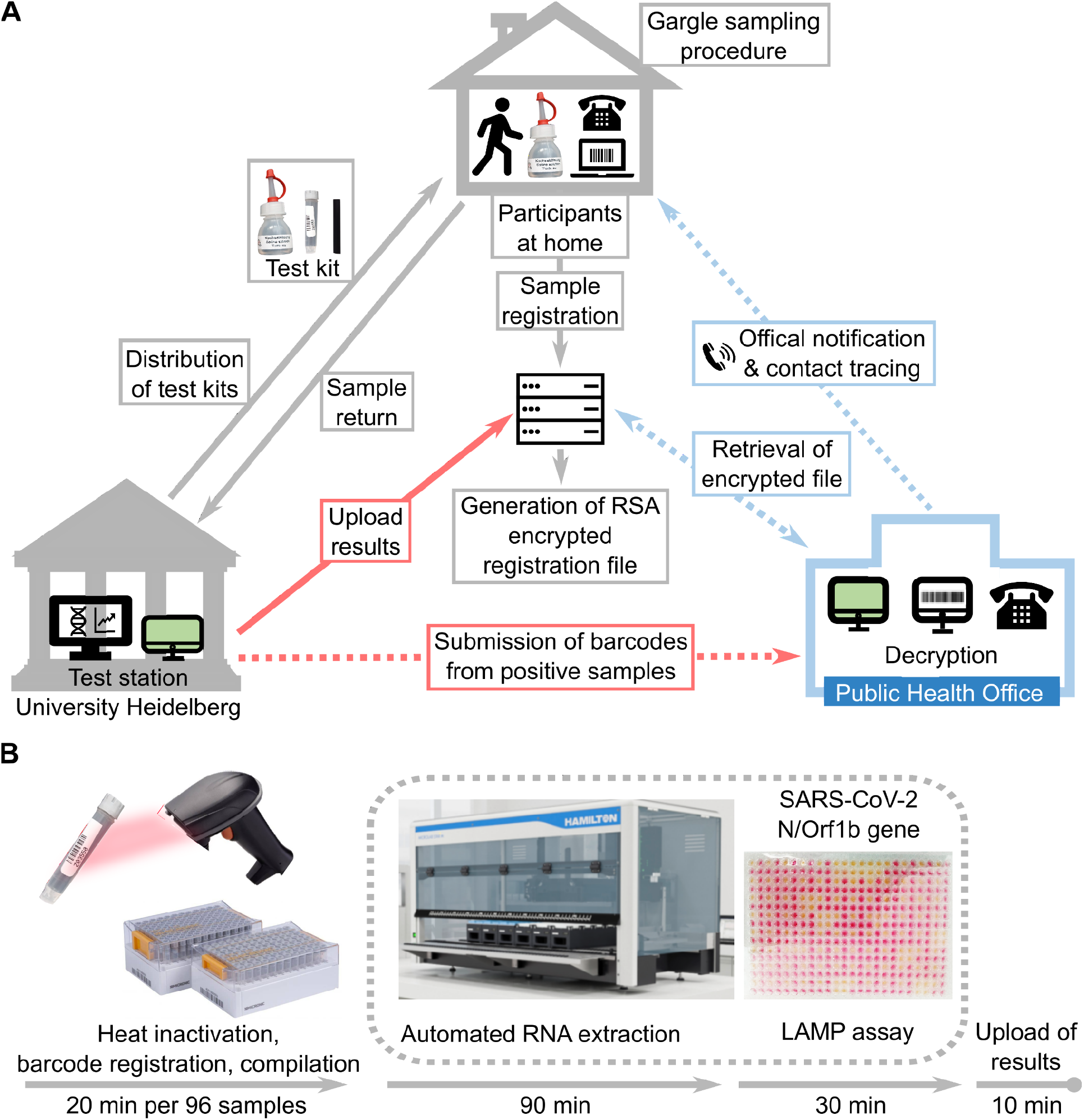
Gargle sample logistics and workflow of high-throughput SARS-CoV-2 diagnostics by RT-LAMP. (**A**) Participants were asked to register their barcoded test tube online via an individual access code previously linked to the test tube. The individual access code was provided with every test kit on a printout. Upon entry, personal information was encrypted with the public key of an RSA key pair and stored using the barcode of the test tube as an identifier of the encrypted contact details. The barcode served as a pseudonym throughout sample analysis. Using a web interface test participants could use the access code to query the status of their sample and the result of the analysis. Upon validation of a putative positive sample by RT-qPCR a report was filed to health authorities that could retrieve the encrypted data file of individual positive samples. The decryption of the data happened at the health authorities that were the only institution in possession of the private key for decrypting the contact information. (**B**) Sample processing and analysis workflow in the test station. The minimum time for each operation without setting up time are shown. When arrived at the test station, samples were heat-inactivated followed by registration, sample plate compilation, RNA extraction, and RT-LAMP analysis. RT-LAMP results were evaluated and finally results were posted online.

### Campus-wide surveillance testing and RT-LAMP sequencing

SARS-CoV-2 surveillance testing started in July 2020 first involving participants from one institute (∼230 employees) during a period of low incidence (<10 cases per week per 100,000 people). We performed 100-200 tests per week initially using mechanical 96-channel pipetting devices (Liquidator 20 μl and 200 ul, Mettler Toledo) for RNA isolation and RT-LAMP assays. In November 2020, a liquid-handling workstation (Hamilton NGS STAR) was installed and the automated RNA isolation protocol was established and validated (Supplemental Figure S2). In the following months, the number of participants (institutes, administrators, and students) increased, resulting in more than 1,500 samples per week in May 2021 (Figure 5A). With the end of the third wave, participation dropped again. At the end of the year (November 23rd to December 22nd, 2020), we additionally analyzed more than 7,000 samples that were part of a surveillance study conducted by the Global Health Institute of Heidelberg (36). Until the end of December 2020, we identified in total 44 SARS-CoV-2 positive samples out of 12,100 analyzed samples (0.36 %) which was higher than what could have been expected for unbiased surveillance testing in a period where the incidence never crossed 0.2 % cases per week (Deckert *et al*., submitted). RT-LAMP-positive samples were validated using a diagnostic RT-qPCR, which yielded Ct values in the range of 17 up to 36. For three samples with only one RT-LAMP positive replicate, a Ct=40 was observed by RT-qPCR, which obtained a diagnostic result of ‘weakly positive’, in agreement with the policy of the clinical diagnostic at that time (December 2020). It should be noted that during the study, the diagnostic interpretation of weakly positive RT-qPCR test results changed. Initially, results in the range Ct35-40 were diagnosed as very weak positive and often reported as positive to the authorities, depending on the RT-qPCR test reagents used, which were changed more often during this period (due to limited supply). Later, from March 2021 on, samples with Ct>37 and from February 2022 on samples with Ct>35 were diagnosed as negative.

**Fig. 5:**
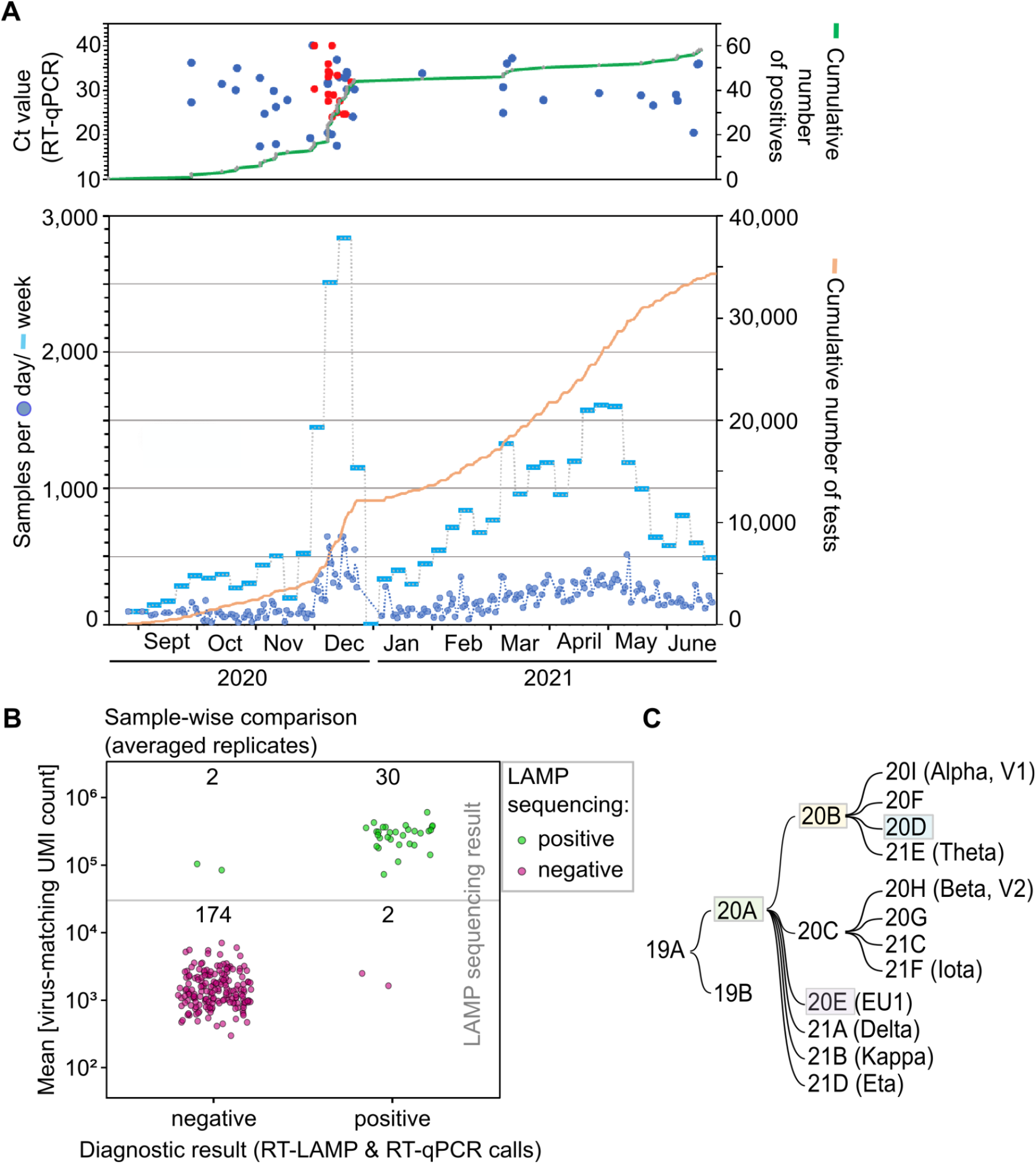
Surveillance testing using the RT-LAMP test facility and validation of results using LAMP-sequencing. (**A**) Time course of surveillance testing (end of August 2020 to end of June 2021). The upper panel shows positive samples only, the lower panel shows the numbers of all tests performed. (**B**) Evaluation of the diagnostic results by LAMP-sequencing. Samples were classified into positive and negative based on the results of three RT-LAMP replicates and a confirmatory RT-qPCR. Of all study samples, a subset of 208 samples was selected, 32 samples with positive diagnostic results). The LAMP sequencing result is shown as the mean UMI count of virus-matching sequences across the three RT-LAMP replicates for each sample. Based on a cutoff of 30,000 UMIs (gray line) the samples are categorized into LAMP-sequencing positive (green) and negative (red). The numbers of samples in each category are indicated. (**C**) Phylogenetic relationships of SARS-CoV-2 clades, as defined by Nextstrain. Variants found in our study were assigned to the highlighted clades using viral whole-genome sequencing.

We (24,26) and others (25) developed LAMP-sequencing as an alternative evaluation method for RT-LAMP results, which enables the analysis of many samples in parallel using a DNA barcoding strategy. In a 96-well plate format using a 96×96 barcoding complexity (96 barcoded Tn5 adaptors and 8×12 barcoded PCR primers) up to almost 10,000 samples (96^2^) can be analyzed in parallel. Since LAMP sequencing is based on the detection of viral sequences not present in the LAMP primers it allows the discrimination of true positives from false-positive RT-LAMP results that occur due to spurious reaction products emerging in many samples 40 min after the start of the RT-LAMP reaction using the described LAMP primer sets. We validated a total of 208 samples by LAMP-sequencing, including 32 samples with positive diagnostic results and 176 samples with negative diagnostic results. By using the mean Unique Molecular Identifier (UMI) count of virus-matching sequences as determined by LAMP-sequencing across the three RT-LAMP replicates for each sample, 30 positive samples and 174 negative samples were confirmed as true positive and true negative, respectively (Fig 5B). We observed that two of the three samples with a weak LAMP positive result and a clinical Ct=40 turned out to be LAMP-sequencing negative, while one sample turned out to be positive. In contrast, two RT-LAMP positive samples that did not yield a positive RT-qPCR result turned out to be positive by RT-LAMP-sequencing. These results indicate a low false-positive rate in our surveillance testing strategy and also demonstrate that there are intrinsic technical limitations for both methods RT-qPCR and RT-LAMP when it comes to the exact diagnosis of samples with extremely weak viral load.

A further important aspect of pandemic control is the detection of the exact viral lineage to trace the spread of new emerging variants. Using isolated RNAs from positive samples and the ARTIC PCR-tiling amplicon protocol and Nanopore sequencing (37), we obtained whole-genome sequences of samples with a sufficient viral load (∼ Ct<32) and detected mutations typical of four major SARS-CoV-2 lineages, namely 20A, 20E, 20B, and 20D (Fig 5C, Supplementary Figure S5).

Taken together, we established scalable surveillance testing and demonstrated the identification of positive samples with a frequency above the positive rate expected from the incidence at the time. Our results demonstrate that the implemented RT-LAMP test station in combination with RT-qPCR for validation of results and LAMP-sequencing for long-term monitoring of test performance provides exceptional sensitivity and specificity to viral testing at the limit of what is currently technically possible while maintaining independence on dedicated and in pandemic situations potentially limited diagnostics supply. Viral genome sequencing in addition enables direct monitoring of the epidemiological dynamics of virus spreading.

## DISCUSSION

### Implementation of a large-scale RT-LAMP-based SARS-CoV-2 testing infrastructure for infection surveillance

Here we describe the development of RT-LAMP-based diagnostics for SARS-CoV-2 surveillance testing in an academic setting suitable for sensitive surveillance and sentinel testing in a low-incidence period of the pandemic. Since the onset of the pandemic, a large number of publications emerged describing the use and validation of RT-LAMP assays for SARS-CoV-2 diagnostics, i.e. (20,27,38–44). These publications describe many different primer sets and use patient samples without purification (“direct assays”) (some are summarized here: (42)) or with prior purification of RNA, many with optimized procedures where compelling data could be obtained that suggest suitable sensitivity of RT-LAMP for detection of infections in asymptomatic and symptomatic subjects. Yet, intrinsic limitations of simple RT-LAMP assays using colorimetric readouts without sequence-specific probes such as molecular beacons, complicate its practical implementation. Here we demonstrate the practicality of RT-LAMP for large-scale surveillance testing over an extended period, thus showing that RT-LAMP is a sensitive, specific, and cost-efficient alternative for SARS-CoV-2 testing.

For the implementation of the RT-LAMP assay we used NEB enzymes and a colorimetric-based readout, however, royalty-free self-purified Bst-LF polymerase and the HIV-RT have been also reported to work very well for RT-LAMP for SARS-CoV-2 (23). Using self-produced Bst-LF polymerase and HIV-RT preparations we found that slight modifications of the sample analysis procedures (using a fluorescent readout, e.g. (45)) ensure similarly high sensitivity as compared to assays performed with the NEB enzyme mixtures. One advantage of the commercial mixtures is that they include reversible heat-inactivated oligonucleotide inhibitors for the enzymes. This makes it unnecessary to keep the sample mixtures strictly at 0°C before the start of the LAMP reaction.

Regular operation of the test station was done using trained student assistants from scientific study programs who worked 3-6 h during 2-3 days per week, making such a testing platform easy to implement in an academic setting. A scientist was available for monitoring and problem solving, and a physician for confirmation of the final diagnoses.

Taken together, our procedures result in an efficient and cost-effective protocol that combines facilitated sample collection - easily performed at home (Roehr *et al*., in preparation (“From Disgusting and Complicated to Simple and Brilliant: Implementation Perspectives Among Users and Rejectors of Mail-in SARS-CoV-2 Gargle Tests”)) - with simple sample handling in the laboratory and high diagnostic reliability.

### Possibilities for quality control and upscaling of RT-LAMP-based testing

Another advantage of our workflow is that reserve samples are available, which contrasts with direct assays where the entire sample is processed for RT-LAMP analysis (25). When using the test platform for surveillance testing, reserve samples are very important. In surveillance testing, even very low false-positive rates of a diagnostic test can be a problem as they can significantly affect the measured incidence (13,46). To overcome this problem all LAMP-positive samples were additionally validated with RT-qPCR, before a definitive diagnosis was made. By using primers for RT-qPCR that bind outside of the RT-LAMP amplified regions, contaminations from RT-LAMP amplicons were excluded, and an independent result using a method with similar sensitivity was generated. If a test platform is used for contact tracing and diagnosis of symptomatic patients, much higher positive rates can be expected. In that case, a low false-positive rate does not matter. Then occasional RT-qPCR validation of randomly selected positive samples is sufficient to monitor RT-LAMP test performance.

An important matter of frequent testing is the costs. RT-qPCR testing costs remained high in many countries throughout the pandemic, and thus do not permit surveillance testing unless pooling strategies are employed. Such strategies, however, are associated with different trade-offs. Pooling could involve the joint extraction of multiple swabs in a small volume to prevent dilution of the sample and preserve the sensitivity of detection. However, in the case of a positive pool, all individuals in this pool have to be retested to identify the infected individual. This significantly slows down the time-to-result and limits the application of such pooling strategies to confined groups and periods of low incidence, but provides a very efficient and cost-effective testing strategy for specific situations, e.g. in schools (47). RT-qPCR testing offered by commercial providers usually reports back results after 24 – 72 hours, thus making such testing almost useless for surveillance testing. Our results were available approx. 5-6 hours after samples were processed for loading onto the robot, and we routinely started with processing at 10 am, and reported the results between 2-4 pm, depending on the number of samples processed, which could be up to 4 times 92 samples per run. Subsequent runs, if required, could be started approx. 2 hours after the previous run, which would permit processing of 12×8×92=4416 samples per day. To further increase the throughput, we also tested and validated a mini-pooling strategy where 2 or 4 samples were pooled before RNA extraction. With this, doubling or quadruplication of the capacity of the test station is possible, while additionally lowering the costs and keeping reserve samples to identify the positive sample in the mini pool.

For such high throughput applications, our implementation of RT-LAMP analysis using a simple plate scanner to monitor the LAMP reaction and simple PCR machines without integrated color or fluorescence readout, becomes limiting, since this requires that the plates are manually loaded onto the plate reader at the given time intervals. In this high throughput scenario, one could either substitute the time-course measurements with a single endpoint measurement. In fact, only one-time point of the LAMP reaction was used for the automated decision-making by the “LAMP plate viewer tool”. Only in unclear cases, the data of the entire LAMP reaction was inspected for interpretation. Alternatively, if the time to result is less critical, our LAMP-sequencing protocol could be used to assess the results of testing of several thousand samples simultaneously (24,26). This comes with the caveats of the time required for library preparation and the sequencing run. This makes LAMP-sequencing an ideal method for large sample volumes, e.g. for surveillance testing of large populations. We never reached sufficient sample numbers to justify RT-LAMP sequencing analysis, but we originally considered validating all RT-LAMP reactions in the period July-December 2020 using LAMP-sequencing. However, due to storage space limitations and the regulations governing the storage of clinical samples, we had to dispose most samples during the study and thus analyzed only a small set of samples.

This publication is intended to serve as a blueprint for the rapid build-up of alternative high-throughput testing infrastructures as a tool for pandemic preparedness. It provides all standard operating procedures (SOPs) for all required steps from self-sampling to secured result reporting. Routine operation of such an RT-LAMP test station with student assistants is comparatively inexpensive and can likely be maintained in many different academic institutions, with the advantage of scalable test capacity to increase the number of tests, e.g., if a future SARS-CoV-2 mutant or other pathogens require it.

## MATERIALS AND METHODS

### Study design

The study is designed as a prospective sample collection for the development of a novel procedure using RT-LAMP for rapid diagnosis of viral infections in a larger population. The present study is exploratory. New samples were collected voluntarily. Detailed information material about the study was prepared for the subjects. Subjects had electronic access to this information material only (study website on the Internet). All subjects provided consent to participate in the study electronically. The subjects had all the information available to make a decision. Further information could be requested by email or telephone.

### Sample handling

Samples were obtained as gargle fluid in 0.9% NaCl solution (B.Braun, Melsungen, Germany) from students and employees at Heidelberg University. Samples were collected in 1.4 ml sterile screw-cap tubes (Micronic, Lelystad, The Netherlands) and taken to the test station within a few hours. Samples were either stirred in a glass beaker with hot water (85-95°C) for 5 min before immediate analysis (to inactivate the samples (24)), or stored at 4°C until further processing. Barcodes on the sample tubes were scanned and registered on our web server. Results were uploaded onto the web server (https://coronatest-hd.de/en/results) and could be retrieved by the participants with access codes linked to the barcodes on individual test tubes. Positive results were validated with diagnostic RT-qPCR and reported to the health department for contact tracing. Samples were processed in a biosafety level 2 (BSL-2) laboratory according to standard microbiological and diagnostic practices.

### Ethical approval

For test development and validation, the trial was approved by the ethics committee of the Medical Faculty at the University of Heidelberg file numbers S-148/2020 and S-392/2020. For the “Virusfinder” study, the trial was approved by the ethics committee of the Medical Faculty at the University of Heidelberg on 02/11/2020, amendment 09/11/2020, file number S-790/2020. The study was conducted in accordance with the Declaration of Helsinki ethical standards. Participants provided consent online via the test station web site before sample and data collection.

### Longitudinal sample collection

Longitudinal samples were obtained from scientist probands and colleagues who had caught Covid-19. The longitudinal positive gargle samples were collected for (i) evaluation of the stability of gargle samples under different storage conditions and (ii) evaluation of factors that could affect the viral load in gargle samples. These samples were kindly provided by some of the participants who were tested SARS-CoV-2 positive and were collected at different time points, before and after specific behaviors (such as tooth brushing, eating, etc).

### Positive and negative controls

Positive controls were diluted from a strongly positive gargle sample (with CT value ∼20) kindly provided by a positive participant. The sample was diluted in 0.9% NaCl solution to achieve the indicated Ct values (Ct29, Ct32, Ct36; corresponding to approximately 8 × 10^4^, 10^4^, and 625 genome copies/ml samples respectively). 0.9% NaCl solution served as negative control.

### RNA isolation with magnetic beads

After heat inactivation and sample registration by barcode scanning, RNA was isolated by an automated liquid handler (Microlab STAR, Hamilton, Bonaduz, Switzerland) with SiMAG-N-DNA magnetic beads (Chemicell, Berlin, Germany). The layout of the instrument desk was designed to allow the processing of up to four 96-position sample racks with micronic tubes at a time. After selecting the program according to the number of samples, a corresponding amount of 2 ml deep-well plates (DWP) were loaded at shown positions in the dialog box. Four buffer troughs were then loaded at the required positions in the dialog box. Corresponding volumes of lysis buffer (containing 5 M guanidinium thiocyanate, 25 mM sodium citrate, 40 mM dithiothreitol, 20 μg/ml glycogen, 1% Triton X-100, and an internal control RNA spiked into the sample lysis buffer before RNA isolation (2000 genome copies/sample)), solution with magnetic beads, 70% ethanol, and nuclease-free water were filled in the troughs according to instructions. After loading different types of tips, 96-well PCR plates for RNA elution and sample racks were loaded at the designated positions. Once started, the instrument automatically ran through all the following steps for RNA isolation: distribution of 140 μl lysis buffer into a defined number of DWPs; pipetting of 140 μl sample into the DWP and mix thoroughly; distribution of 200 μl of magnetic beads solution into the DWP containing the sample lysates and shake the DWP at 1200 rpm for 15 min; placing the DWP on a magnet plate for 5 min; discarding supernatants and wash the magnetic beads three times with 200 μl 70% ethanol; rinsing the magnetic beads briefly with 100 μl nuclease-free water; resuspension of the magnetic beads in 60 μl nuclease-free water; transfer of the eluted RNA into corresponding 96-well PCR plates.

### RT-LAMP assay

The RT-LAMP assay was conducted in 384-well PCR plates. Each sample was analyzed in three technical replicates with N2/ORF1b-2 primers for detection of SARS-CoV-2 RNA, and one replicate with specific primers for detection of the spike-in RNA (internal control). The Master mix was prepared at room temperature immediately before use. Per 12.5 μl reaction, the Master mix contained 6.25 μl Warm Start Colorimetric LAMP 2x Master Mix (M1800, New England Biolabs), 0.125 μl 100x primer mix, 0.5 μl 1 M guanidine hydrochloride, and 0.625 μl RNase-free water. The master mix (7.5 μl per reaction) was aliquot into a 384-well plate, and 5 μl of RNA was added into each well and mixed thoroughly. Plates were then sealed with a transparent adhesive foil (MSB1001, Bio-Rad), and incubated in a 384-well thermal cycler (Biometra) at 65 °C for in total 30 min with the lid heated to 75 °C. During the incubation, plates were removed from the thermal cycler at indicated time points (0, 10, 15, 20, and 30 min), and placed on an ice-cold metal block for 1 min to pause the reaction. The absorbances of each well at 434 and 560 nm were measured with the Tecan Microplate Reader (Tecan Trading AG, Switzerland). To quantitatively indicate the colorimetric change during the acidification of the LAMP reaction (434 nm absorbance increased, 560 nm absorbance decreased), delta OD value (ΔOD) was defined as the difference between absorbances of phenol red at two different wavelengths: ΔOD = OD_434nm_ - OD_560nm_.

### RT-qPCR

Master mix was prepared with 4.0 μl LightCycler® Multiplex RNA Virus Master (Roche, Basel, Switzerland), 0.5 μl LightMix® Modular SARS-CoV (COVID19) *E* gene (cat. no. 53-0776-96, TIB MOLBIOL Syntheselabor, Berlin, Germany), 0.1 μl reverse transcriptase enzyme (Roche, Basel, Switzerland), and 5.4 μl RNase-free water per reaction. After distributing 10 μl of master mix to a 96-well plate, 10 μl of RNA were added to each well. A positive control for the *E* gene was used to validate the performance of the RT-qPCR. The RT-qPCR was running in the following condition: reverse transcription at 55 °C for 5 min, denature at 95 °C for 5 min, followed by 45 cycles of amplification at 95 °C for 5 sec, 60 °C for 15 sec, 72 °C for 15 sec, and then cooling at 40 °C for 30 sec. A total of 10^3^ genome copies of E gene RNA per RT-qPCR reaction is equivalent to a cycle threshold (Ct) value ≈ 30.

### LAMP sequencing

The three technical replicates of each sample after RT-LAMP reaction were individually processed and analyzed by LAMP-sequencing. LAMP-sequencing was performed and analyzed as previously described [DaoThi’20, Herbst’21] except using primer P7nxt-N2-Lbrc (sequence 5’-GTCTCGTGGGCTCGGAGATGTGTATAAGAGACAGGGTCAACCACGTTCCCGAAG-3 ‘) for NGS library preparation to accommodate that the RT-LAMP reactions were performed with the N2 primer set. A LAMP-sequencing read was considered as virus-matching if one of the following sequences was detected (in 5’-3’ direction): CCAGCGCTTCAGCG, CTTCGGAATGTCG, TGTCGCGCATTGG, TCACACCTTCGG. LAMP-sequencing samples were considered positive if more than 30,000 virus-matching UMIs were observed.

### Whole-genome sequencing of SARS-CoV-2 samples

SARS-CoV-2 genomes were sequenced using the ARTIC long-amplicon tiling multiplex method based on Oxford Nanopore Technologies (ONT) sequencing, according to the protocol reported in (https://www.protocols.io/view/ncov-2019-sequencing-protocol-v3-locost-bp2l6n26rgqe/v3, version by Josh Quick), with the improvements described in (37) and the nCoV-2019 PCR primer set V3 (https://github.com/artic-network/artic-ncov2019/blob/master/primer_schemes/nCoV-2019/V3/n

CoV-2019.tsv). Half-scale PCR reactions were used (11.25 μL each) to save costs, and 1.25 μL cDNA was added to each reaction as a template. We used 25 cycles of amplification for all samples, and we decreased the annealing/extension temperature to 63°C, since we observed dropout of amplicon 64 in our preliminary experiments. The ONT Native Barcoding Expansion Kits 1-12 (EXP-NBD104) and 13-24 (EXP-NBD114) were used for barcoding different samples (always including negative controls), and the pooled libraries were loaded on R9.4.1 flow cells (ONT) for multiplexed sequencing on a MinION device (ONT).

### Bioinformatic analysis of SARS-CoV-2 whole-genome sequencing data

The Guppy basecalling software (v4.4.2; ONT) with the high-accuracy mode was used for basecalling Nanopore sequencing data. Demultiplexing and barcode trimming were performed using the “guppy_barcoder” function (v4.4.2; ONT). This was followed by filtering using the “artic guppyplex” function of the ARTIC pipeline for SARS-CoV-2 ONT sequencing (https://github.com/artic-network/artic-ncov2019), which removed reads with quality (Phred) scores below 7 and retained only reads of length between 400 and 700 bp to exclude chimeric reads. The tool NanoPlot (https://github.com/wdecoster/NanoPlot) was used for quality control of sequencing data. Downstream analyses and generation of consensus sequences were performed with the “artic minion” workflow of the ARTIC pipeline, using “minimap2” to align reads to the reference genome sequence, and “nanopolish” to call variants. Any position of the reference sequence that was not covered by at least 20 reads was considered of insufficient coverage during generation of the consensus sequence. Consensus sequences of all samples were further analyzed using Nextclade CLI (v1.0.0; https://github.com/nextstrain/nextclade), the list of signature mutations defining the Nextstrain clades (update of June 3, 2021; https://github.com/nextstrain/ncov/blob/master/defaults/clades.tsv), and a representative set of 1849 SARS-CoV-2 genome sequences from the GISAID database (https://www.gisaid.org) for phylogenetic classification (in Nextclade v1.0.0, last updated on June 16, 2021). We used the genome sequence of SARS-CoV-2 isolate Wuhan-Hu-1 (GenBank accession number: NC_045512) as the reference sequence in our analyses. Results were further analyzed and plotted using R (R Core Team. R: A language and environment for statistical computing [Internet]. Vienna: R Foundation for Statistical Computing; 2020. [cited 2020 Sep 11]. Available from: URL https://www.R-project.org/).

### RT-LAMP evaluation software

LAMP plate viewer tool is available on GitHub at https://github.com/anders-biostat/lamp_plate_analysis. The tool consists of three files. The main part is an R (48) script that is used for reading in the data (with the “readxl” (49) and “tidyverse” (50) packages), generating interactive visualization (the “rlc” package), and sending results to the server (the “httr” package (Hadley Wickham (2022). httr: Tools for Working with URLs and HTTP. R package version 1.4.3. https://CRAN.R-project.org/package=httr)). The accompanying HTML file is opened by the “rlc” package and contains a layout for the interactive plots, some

CSS styling, and additional JavaScript functionality. The batch file contains a single “Rscript” command that starts the R script using the path to the plate reader output file as an argument. The script relies on the presence of data files with the barcode assignment to each well and the exact directory structure that was used in the testing facility. The viewer is then opened in a default browser window.

For automatic sample classification, first, a decision is made separately for each well of the 384-well plate. To this end, the maximum ΔOD value within the first 20 min is compared to the preset threshold of 0.1. Every well that produced a maximum ΔOD value above the threshold is considered positive, all other wells are assigned negative status. Also, a baseline is calculated for each well as a mean ΔOD value during the first 10 minutes. A well with a baseline above −0.1 is considered contaminated with magnetic beads after the RNA extraction step. Next, assigned statuses for all the replicates of the given sample are pooled together and the classification is performed as follows (in this order):

- If at least one well has a baseline above −0.1 - “repeat”
- If all wells with the N2 primer are positive - “positive”
- If the spiked-in control is negative - “repeat”
- If all the wells with the N2 primer are negative - “negative”
- Anything else - “inconclusive”

In addition, if the file with barcode assignments contained any comments (usually used to indicate some problems with the sample), the “repeat” status was assigned no matter the results, thus highlighting the sample for manual classification. The “failed” status (indicating that the sample can not be processed) could be only assigned manually. During the study period, the classification algorithm was changed multiple times, based on the needs and observations of the lab personnel. Since the final decision has always been made manually, these adjustments affected the convenience of the users rather than the actual results.

## Data Availability

All data produced in the present work are contained in the manuscript or upon reasonable request to the authors.

## Acknowledgements

We thank the Heidelberg “Coronatest team” for having enabled this study: Manuela De Allegri, Hoa Thi Nguyen, Matthias Sand, Lisa Koeppel, Lena Maier Hein, Tobias Roß, Tim Adler, Tobias Siems, Stephan Brenner, Michael Marx, Freda Röhr, Ferdinand Uellner, Lucia Brugnara, Shannon A. McMahon, Aurelia Souares and Till W. Bärnighausen. We are also very grateful for the support of the Rectorate of Heidelberg University, the various faculties and institutes of Heidelberg University, and many individual laboratory directors who encouraged participation. Additionally, we thank Jörn Bargstedt, Adrien Simon Jolly and Erik Schwabe for the provision of longitudinal samples.

## Author contributions

Conceptualization: MK, SA

Methodology: MK, MM, DL, RB

Software: SO, AU, SA

Validation: RB, DL, MM

Formal analysis: DL, SD, KH, IAP

Investigation: MM, DL, DY, MJ, VW, DU,

Resources: KG

Data Curation:

Writing - Original Draft: MK, DL

Writing - Review & Editing: ADz

Visualization: MK, DL, ADz, IAP, KH,

Supervision: MK

Project administration: DK

Funding acquisition: MK, VLDT, HGK, ADe, CMD

## Data and materials availability

All software is available on Github.

---

**Supplemental Fig. S1:**
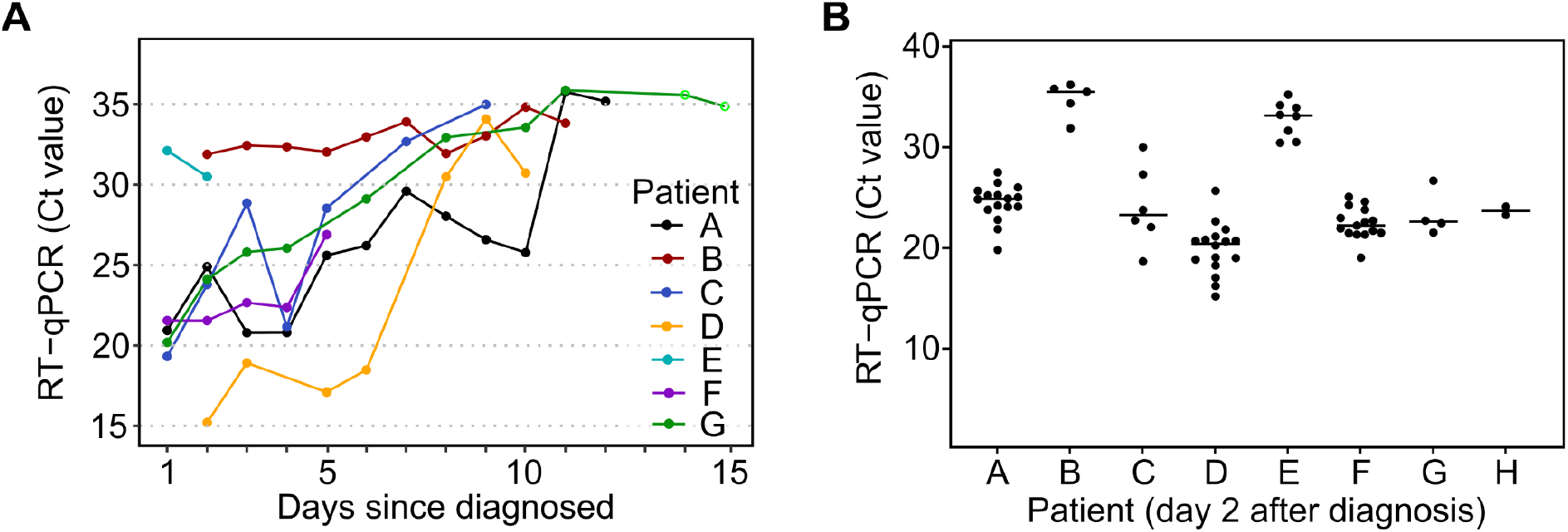
Analysis of longitudinal samples using RT-qPCR. (**A**) Viral loads of early samples (collected immediately after waking up in the morning) on different days since the patient was diagnosed with COVID-19. (**B**) The fluctuations of the viral load within one day. Each dot represents one timepoint (between 7 am and 12 pm).

**Supplemental Fig. S2:**
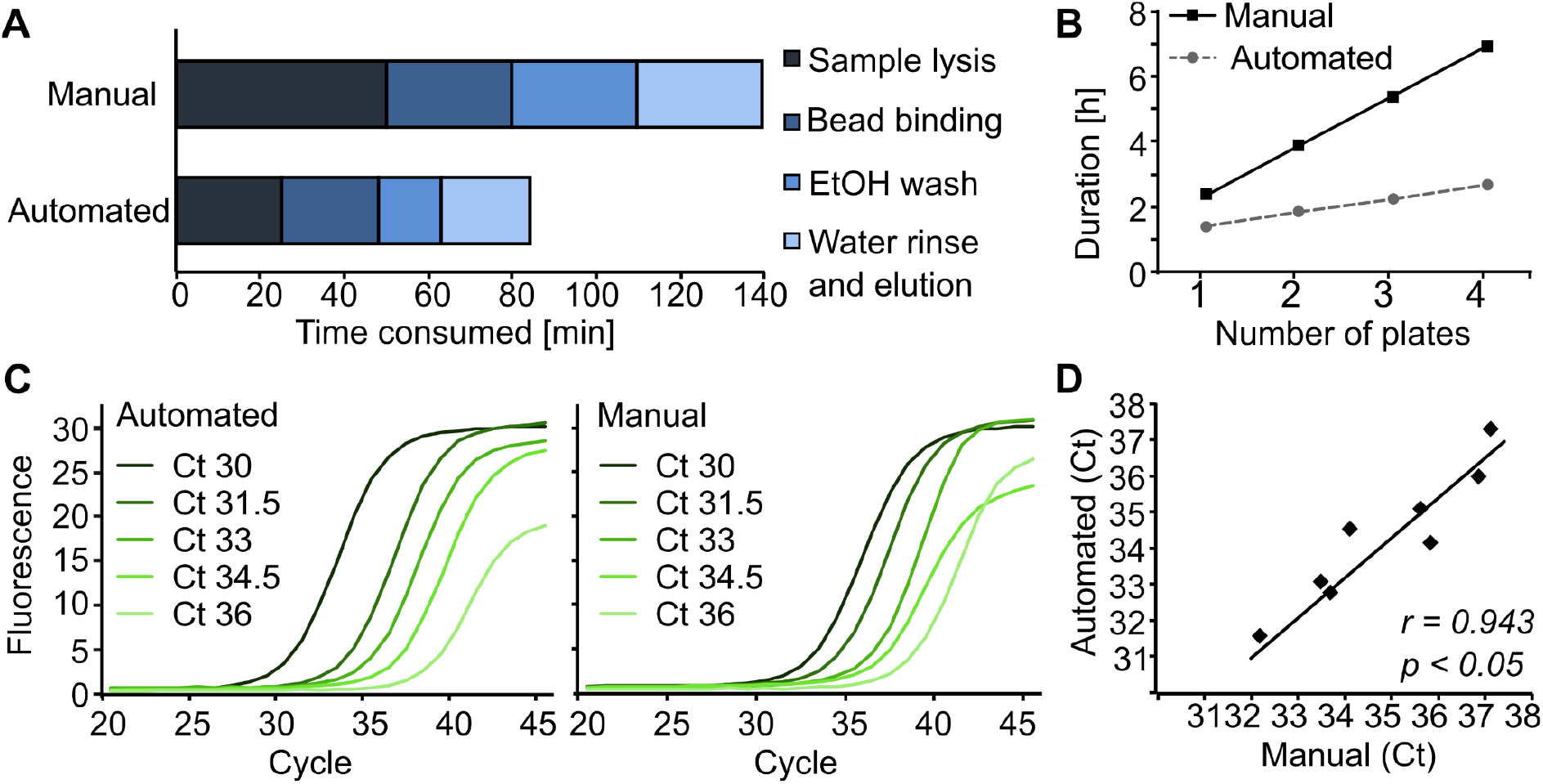
Comparison of manual and automated RNA extraction. (**A**) Timewise comparison of manual and automated RNA extraction from 96 samples. Time consumed by sample lysis, bead binding, wash, and elution is shown in different colors. (**B**) Timewise comparison of manual and automated RNA extraction from multiple 96-well plates. (**C**) RT-qPCR amplification curves of template RNA isolated automatically (left) and manually (right) from serial diluted SARS-CoV-2 positive gargling samples. (**D**) Correlation of Ct values between automatically and manually extracted template RNA.

**Supplemental Fig. S3:**
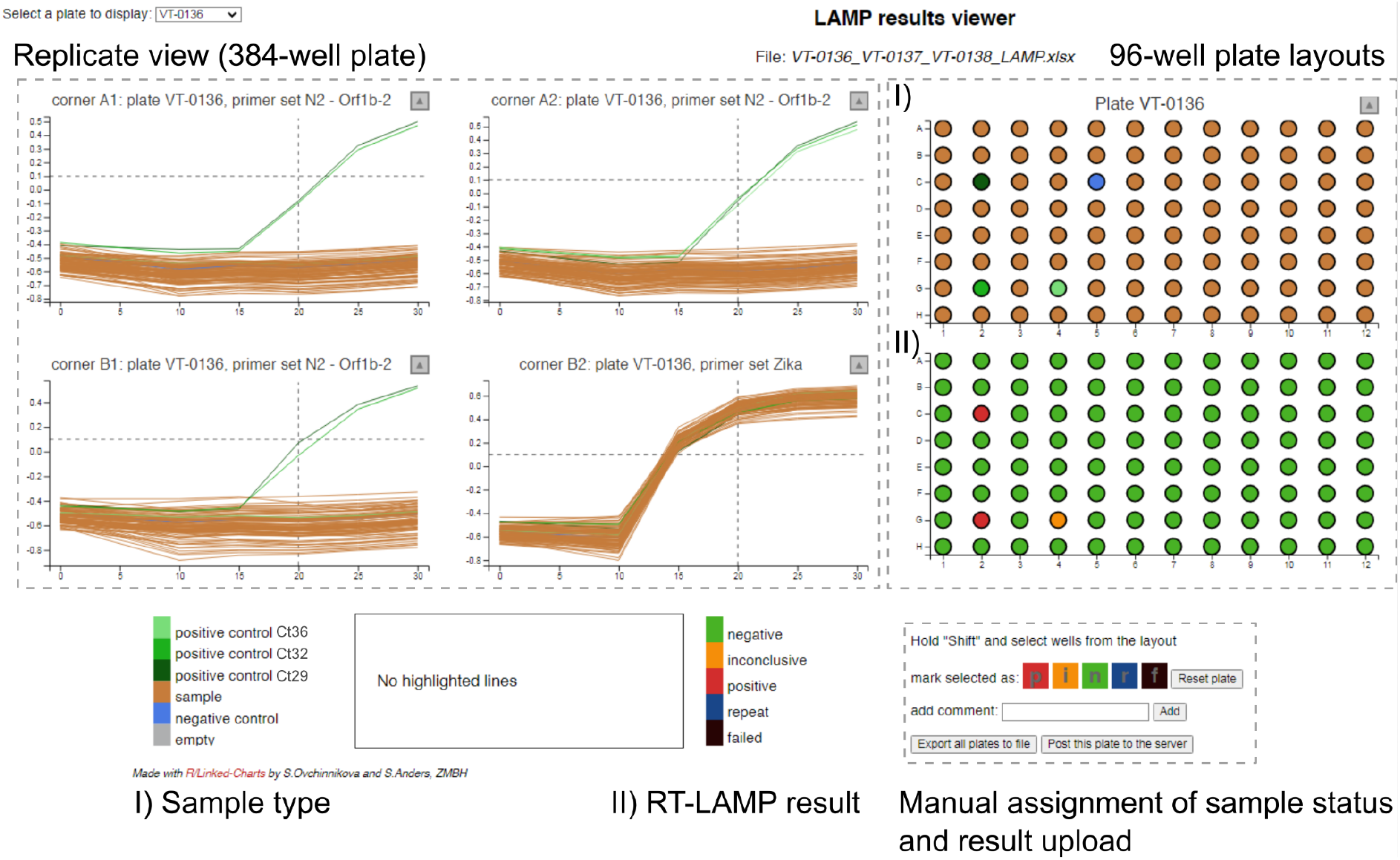
LAMP plate viewer user interface. The plate reader output file is fed into the LAMP plate viewer tool together with the barcode and position information of each sample in the 96-well sample plate. Each sample plate is displayed individually. On the right the 96-well sample plate layouts color-coded according to 1) sample type (test sample, controls, empty wells) or 2) automatically assigned classification of RT-LAMP results (green = negative, red = positive, orange = inconclusive, blue = repeat and black = failed) are shown. On the left, the RT-LAMP measurements for each well in the 384-well plate are visualized (replicate view) to allow quality control by the test station personnel. The tool is interactive and allows to highlight all replicates of a single test sample by selecting the corresponding well in one of the layouts of the 96-well plate on the right. The classification of the RT-LAMP results is done automatically, but the test station personnel can edit the sample statuses if necessary before the results of the sample plate are uploaded to the server.

**Supplemental Fig. S4:**
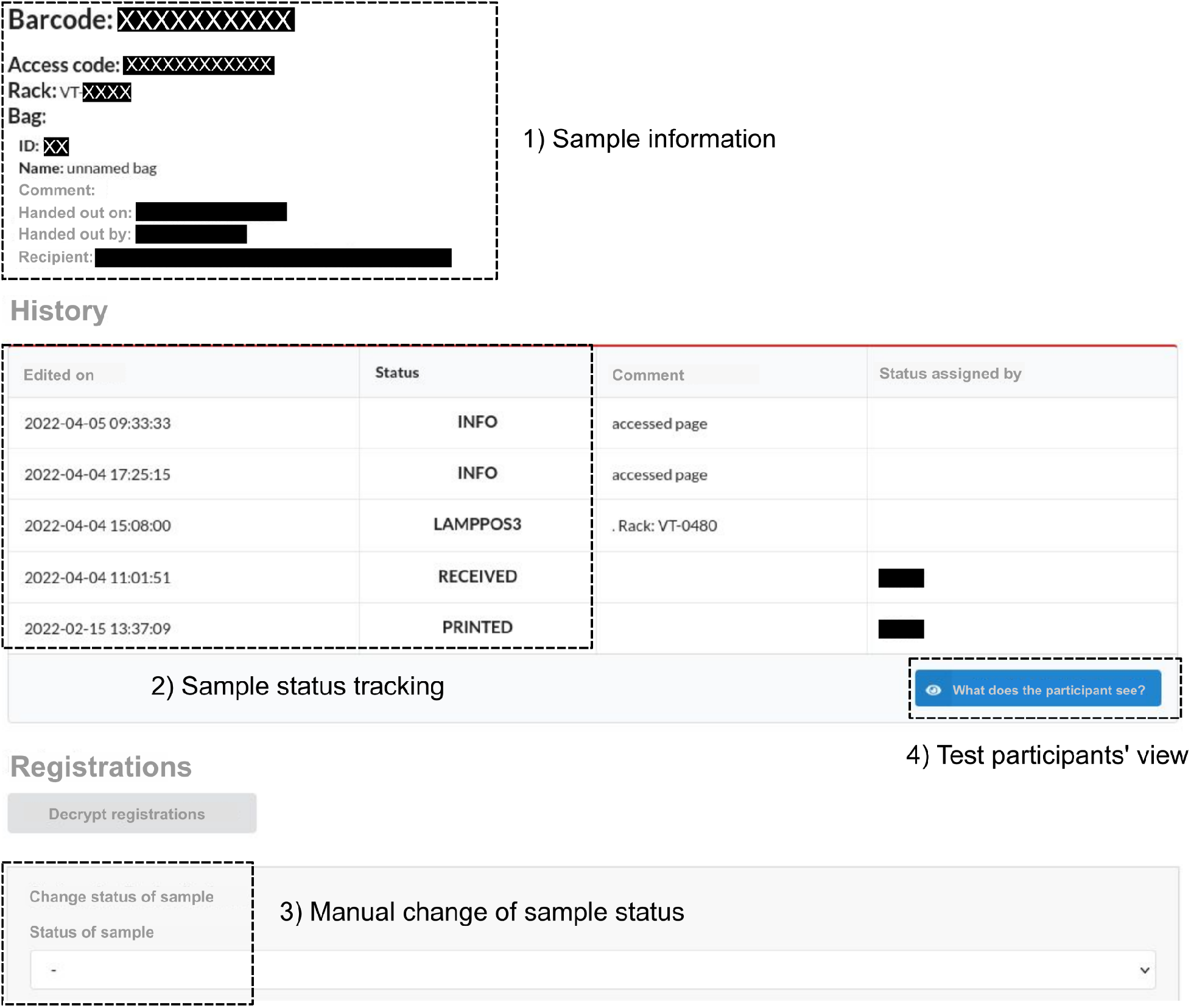
Example entry in the database user interface. The database user interface allowed the test station personnel to track and edit all registered samples. Labels in grey were translated from German into English. 1) Sample information: Samples were identified by their barcode on the Micronic test tube. Additionally, the distribution of test kit bags (‘Abnehmer’) could be documented, e.g. to settle the costs of testing). 2) Sample status tracking: Individual samples were assigned a corresponding status, which was updated in each phase of the workflow. These include the generation of an access code for each Micronic test tube before distribution of the test kit to a participant (PRINTED), sample arrival at the test station (RECEIVED), RT-LAMP results (LAMPNEG versus LAMPPOS or LAMPINC in case of uncertainty) and the result retrieval by the test participant (INFO - accessed page; not visible for the test participant). 3) Manual change of sample status: The test station personnel could edit the sample status, e.g. after verification of a positive result via RT-qPCR (PCRNEG/ PCRPOS). 4) The test participants’ view allowed the test station personnel to view the result page, which was shown to the user with the current status set.

**Supplemental Fig. S5:**
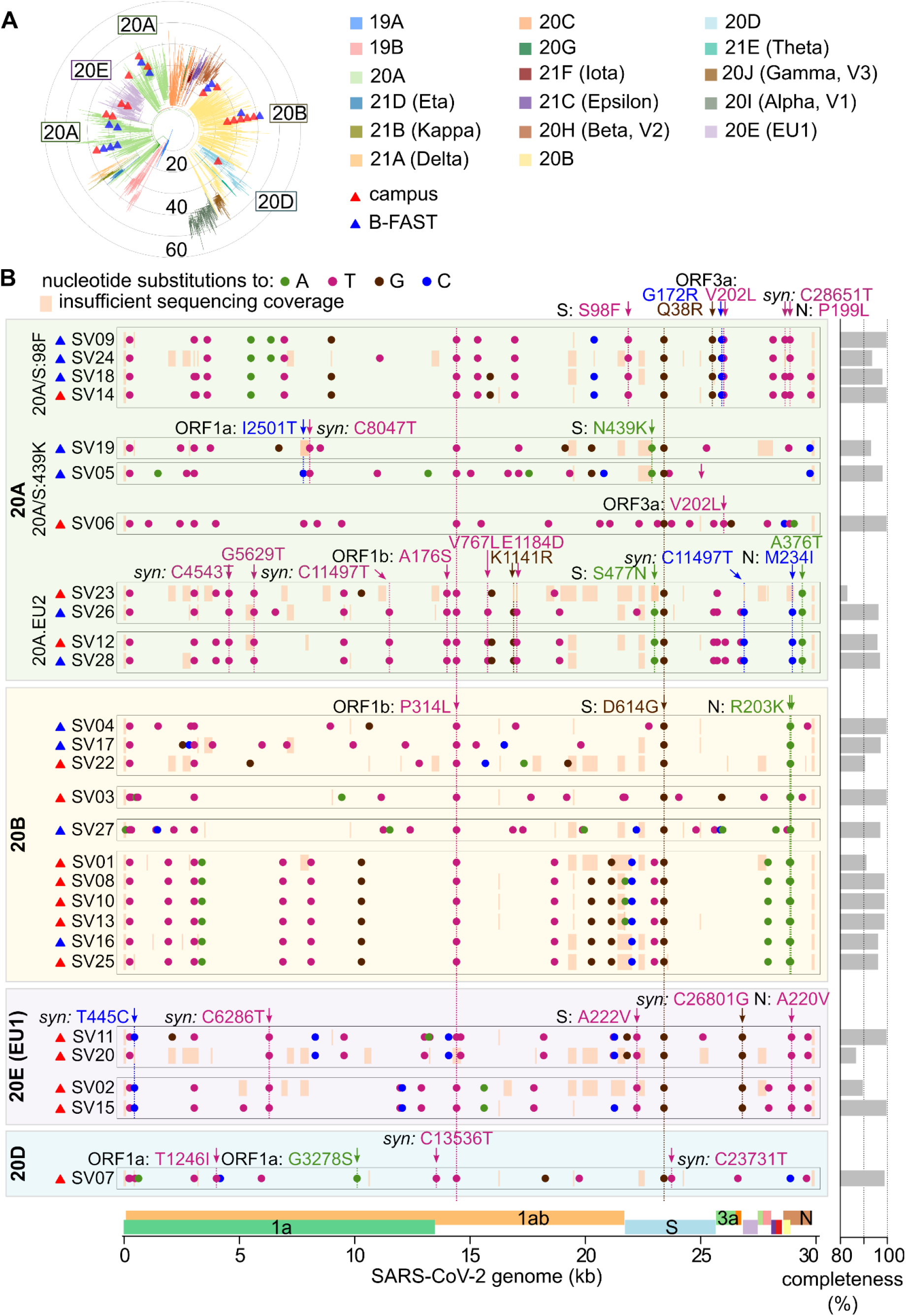
Detailed characterization of sequenced SARS-CoV-2 viral genomes with the detected mutations per sample. **(A)** Phylogenetic placement of the 27 obtained genome sequences (triangles) on the Nextclade tree of 1849 reference SARS-CoV-2 genome sequences (representing the genetic diversity of circulating SARS-CoV-2 at the time of the analysis). **(B)** Mutations of the 27 whole-genome sequences in comparison to the original reference strain (Wuhan-Hu-1). Sequences were grouped and ordered according to their phylogenetic placement. Specific signature mutations that define the various Nextstrain clades are indicated when present in the analyzed samples. Stretches of genomic sequence with insufficient sequencing coverage for reliable mutation calling are indicated by orange boxes. The completeness of each genomic sequence is shown on the right.

**Table S1:** Specificity and sensitivity of the RT-LAMP assay. Sensitivity (for each Ct bin) and specificity of the RT-LAMP test with respect to the RT-qPCR results, as derived from the results shown in Fig. 3.

| Ct range | Sensitivity [%] | 95% CI [%] | Specificity [%] | 95% CI [%] |
| --- | --- | --- | --- | --- |
| 0-25 | 100.0 | 95.9 - 100.0 | 99.0% | 97.6 - 99.6 |
| >25-30 | 100.0 | 95.4 - 100.0 |  |  |
| >30-34 | 100.0 | 93.7 - 100.0 |  |  |
| >34-36 | 83.3 | 61.8 - 94.5 |  |  |

